# When Prescribing Diverges From Published Lactation Compatibility Guidance: A National Claims Analysis

**DOI:** 10.64898/2026.10.19.26365940

**Authors:** Kaytlin Krutsch, Duke Appiah

## Abstract

**Purpose:** Prescribing to lactating women is constrained by limited evidence. Published compatibility classifications provide clinical guidance and a discrete research comparator. How often prescribing diverges from that guidance, and what characteristics are associated with divergence, is unknown at national scale.

**Methods:** Retrospective analysis of prescriptions filled during lactation, identified in the 2012-2014 MarketScan Commercial Claims Database. Prescriptions were classified as concordant (Hale’s lactation risk category L1-L3) or discordant (L4-L5). Multivariable logistic regression identified factors associated with concordance.

**Results:** Of 96,484 new prescriptions, 95,710 (99.2%) were concordant and 774 (0.8%) were discordant. Discordant prescriptions were filled later after delivery (median 59 vs 33 days) and had a day supply three times longer (30 vs 10 days). In multivariable analysis (N=28,732), concordant prescriptions were associated with maternal age under 35 (adjusted odds ratio (aOR)=1.34, 95% confidence interval (CI): 1.02-1.75) and chronic-condition medications (aOR=10.94, 95% CI: 6.47-18.50), and were less likely within 14 days of delivery (aOR=0.53, 95% CI: 0.38-0.73) and with each additional day of supply (aOR=0.96, 95% CI: 0.96-0.96). Compared with family practice, concordance did not differ for obstetric, internal medicine, or emergency medicine prescriptions, but prescriptions from other provider types had lower odds of concordance (aOR=0.57, 95% CI: 0.41-0.79).

**Conclusions:** Prescribing during lactation was overwhelmingly concordant with published compatibility guidance; the rare divergence was patterned by prescription and prescriber characteristics. Because the categories are an expert classification rather than consensus guidance, divergence should not be read as inappropriate prescribing.

## 1. Introduction

Medications are commonly needed during lactation, and decisions about them are made with limited evidence. A 2026 landscape analysis found that approximately 40%-50% of drugs prescribed postpartum had no or only weak evidence within individual pharmacology domains, a larger knowledge gap than was observed in pregnancy or pediatric groups^1^. Among 290 new therapeutic products approved by the US Food and Drug Administration from 2010 through 2019, 48.6% had no lactation safety data and only 2.8% had human lactation data^2^. In the absence of trial data, clinicians rely on secondary classifications that synthesize what pharmacokinetic, case, and observational evidence exists into a compatibility judgment.

Hale’s Lactation Risk Categories provide a widely used classification for this purpose. They assign each monograph to one of five levels from L1 (compatible) to L5 (hazardous), and have been used operationally across a range of study designs and settings: as an exposure variable in a Brazilian birth cohort of 3,988 breastfeeding women^3^; as the classification framework in a prospective Greek cohort of 847 women examining medication intake as a determinant of breastfeeding cessation^4^; in cross-sectional assessment of medicine use among 294 women in Kampala^5^; and most recently as training labels for machine-learning prediction of lactation risk^6^. The categories are published biennially, which permits assessment against the information available at the time a prescription was written. The 2012 categories applied in this study represent an expert classification rather than a consensus guideline; divergence from them is therefore not evidence of inappropriate prescribing. Their value here is as a time-matched, discrete research comparator.

What has not been established is how often prescribing during lactation diverges from published compatibility guidance, or what distinguishes the prescriptions that do. Existing studies have described medication use or medication-related problems in birth cohorts, prospective cohorts, cross-sectional surveys, and specialized pharmacist consultation samples^3-5,7,8^, but these designs do not provide the scale possible for a national US claims-based assessment of concordance by prescription and prescriber characteristics. A recent systematic review found that medication-related breastfeeding discontinuation is recurrent and may often be avoidable, although certainty of evidence was very low^9^.

Using a claims-based method to identify periods of probable lactation in a national commercial population^10^, we characterized prescriptions filled during lactation that diverged from published compatibility categories, and examined which patient, prescription, and prescriber characteristics were associated with divergence.

## 2. Methods

This analysis used methods described previously^10^ to create a cohort of prescriptions filled during lactation. In brief, deliveries were identified in the 2012-2014 Merative MarketScan Commercial Claims Database by delivery-related diagnosis-related group, and lactation was identified using surrogate indicators recorded in billing data: breast pump equipment and replacement supplies, the encounter code for supervision of a lactating mother, mastitis and breast abscess, nipple infection and cracked nipple, engorgement, galactorrhea, and lactation class attendance. A lactation window was assigned from the date of delivery to the date of the last such indicator, and prescriptions filled within that window were classified as filled during lactation. A prescription was counted as new when the claim represented an original fill rather than a refill. The complete phenotype and code lists are provided in that report^10^.

The analytic population is a subset of the cohort previously described^10^. Of the 158,471 new prescriptions filled within a lactation window in that analysis, 96,484 were assigned a lactation risk category and form the descriptive sample here. Of these, 28,732 had complete data on all model covariates and form the analytic sample for the regression analysis; the reduction was driven largely by missing prescriber linkage. No new cohort was assembled and no additional data were obtained.

Medications were matched to lactation risk categories from Hale′s Medications and Mothers′ Milk^11^, using the 2012 edition to correspond with the study period. For combination products the most conservative category was applied. Prescriptions for medications categorized L1 (compatible) through L3 (probably compatible) were classified as concordant; those categorized L4 (possibly hazardous) or L5 (hazardous) were classified as discordant. Lactation data availability (limited data, no data, or assessment unavailable) was drawn from the 2014 edition^12^, because the variable was not reported in the 2012 edition. Analyses were restricted to prescriptions with an assigned lactation risk category. Of 5,621 monographs in the 2012 database, 108 (1.9%) were assigned L4 or L5.

Exposure variables were maternal age at the time of the fill, days between delivery and the fill, census region, the MarketScan maintenance indicator (which classifies a medication by whether its primary use is for acute or chronic conditions), day supply, prescriber type, and place of service. Prescriber type and place of service were reconstructed by cross-referencing the date a prescription was written against outpatient encounters on the same date. The multivariable model included only prescriptions linked to an encounter. The outcome was prescribing concordance at the level of the individual prescription.

Age was dichotomized at 35 years, corresponding to advanced maternal age. Days past delivery was dichotomized at 14 days, distinguishing the acute postpartum period from established lactation. Reference categories for region, maintenance indicator, provider type, and place of service were the categories with the largest sample. Prescriptions with an unknown maintenance indicator were excluded.

Characteristics were compared using chi-square tests for categorical variables, t tests for normally distributed continuous variables, and Wilcoxon rank-sum tests for non-normally distributed continuous variables. Univariable and multivariable logistic regression were used to estimate crude odds ratios (OR) and adjusted odds ratios (aOR) together with their corresponding confidence intervals (CI). The multivariable analyses were restricted to prescriptions with complete data on all covariates (N = 28,732). Because the multivariable model was limited to 28,732 prescriptions with complete covariate data, its findings are considered separately from the descriptive comparisons in the full concordance sample. Analyses were conducted with SAS 9.4 (SAS Institute, Cary, NC).

## 3. Results

Of 96,484 new prescriptions included in the concordance analysis (filled within a lactation window and assigned a lactation risk category), 95,710 (99.2%) were concordant and 774 (0.8%) were discordant. Discordant prescriptions had a later time past delivery, with a median (interquartile range (IQR)) of 59 (25-153) days compared with 33 (8-94) days for concordant prescriptions. Mean age at the time of the fill did not differ. Though there was a statistically significant difference in prescribing concordance by region, the differences were small, ranging from 0.6% discordant in the West to 0.9% in the South (Table 1).

**Table 1.**
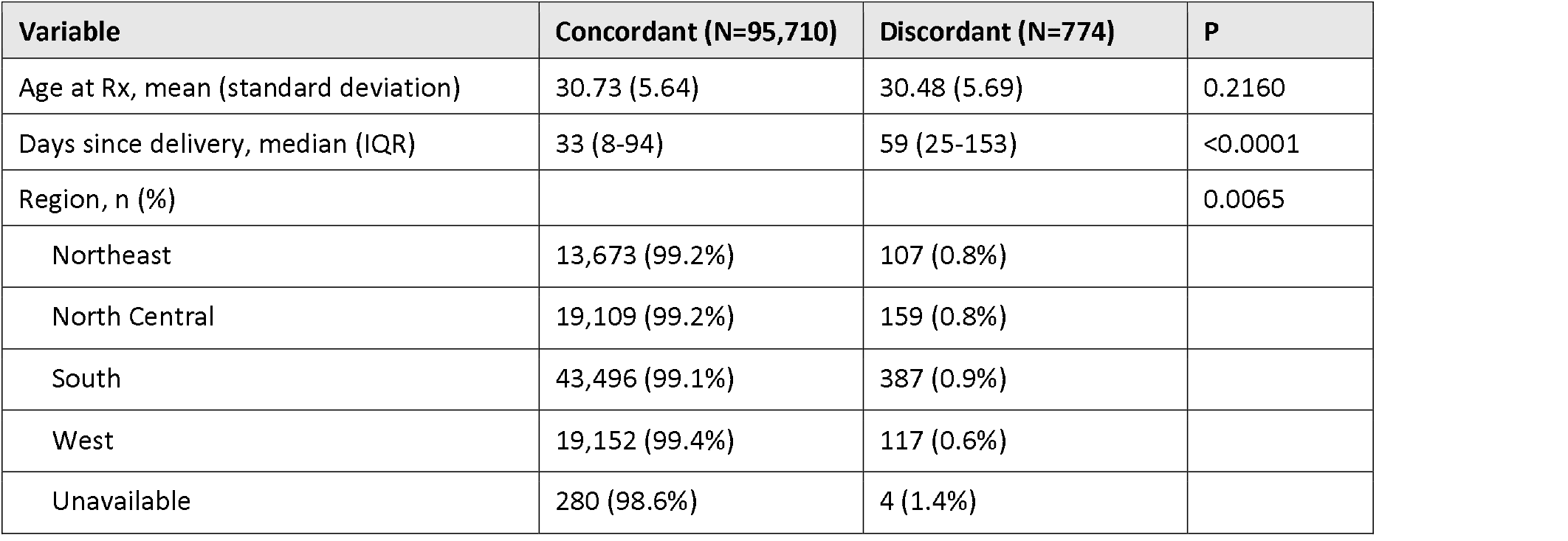
Patient characteristics of new prescriptions filled within a lactation window, by concordance.

Differences in concordance status for prescription characteristics were all statistically significant (Table 2). The median day supply for discordant prescriptions, 30 days (IQR 10-30), was three times longer than that of concordant prescriptions, 10 days (IQR 5-28). Discordance was more common among medications used primarily for chronic conditions (1.6%) than for acute conditions (0.7%). Discordant prescriptions were almost entirely confined to medications for which limited lactation data were available (1.3%) rather than none (<0.1%) (Table 2).

**Table 2.** Prescription characteristics by concordance.

| Variable | Concordant (N=95,710) | Discordant (N=774) | P |
| --- | --- | --- | --- |
| Day supply, median (interquartile range) | 10 (5-28) | 30 (10-30) | <0.0001 |
| Maintenance indicator, n (%) |  |  | <0.0001 |
| Chronic conditions | 10,446 (98.4%) | 175 (1.6%) |  |
| Chronic and acute conditions | 20,845 (99.4%) | 130 (0.6%) |  |
| Acute conditions | 63,982 (99.3%) | 467 (0.7%) |  |
| Other/unavailable | 435 (99.5%) | 2 (0.5%) |  |
| Lactation data availability, n (%) |  |  | <0.0001 |
| Limited lactation data | 58,245 (98.7%) | 758 (1.3%) |  |
| No lactation data | 37,183 (100.0%) | 3 (0.0%) |  |
| Assessment unavailable | 282 (95.6%) | 13 (4.4%) |  |

Discordance was least common among prescriptions linked to obstetric and perinatal providers (0.6%) and family practice (0.7%), and most common among prescriptions linked to other or unknown provider types (1.0%). Prescriptions written at inpatient hospital visits were less often discordant (0.4%) than those at office or clinic visits (0.8%) (Table 3).

**Table 3.** Prescriber characteristics by concordance.

| Variable | Concordant (N=95,710) | Discordant (N=774) | P |
| --- | --- | --- | --- |
| Specialty |  |  | <0.0001 |
| OBGyn/perinatal medicine | 25,686 (99.4%) | 158 (0.6%) |  |
| Family practice | 21,915 (99.3%) | 148 (0.7%) |  |
| Internal medicine (NEC) | 7,349 (99.2%) | 63 (0.8%) |  |
| Emergency medicine | 4,446 (99.1%) | 41 (0.9%) |  |
| Other/unknown | 36,314 (99.0%) | 364 (1.0%) |  |
| Prescriber Location |  |  | <0.0001 |
| Office/clinic | 74,884 (99.2%) | 636 (0.8%) |  |
| Outpatient | 9,155 (99.2%) | 72 (0.8%) |  |
| Inpatient hospital | 5,844 (99.6%) | 25 (0.4%) |  |
| Other/unknown | 5,827 (99.3%) | 41 (0.7%) |  |

In the multivariable analyses (N = 28,732), prescriptions filled for women under 35 had higher adjusted odds of concordance (aOR=1.34, 95% CI: 1.02-1.75). Prescriptions filled within 14 days of delivery had lower odds of concordant prescriptions (aOR=0.53, 95% CI: 0.38-0.73). Compared with the South, the West (aOR=1.50, 95% CI: 1.12-2.02) and North Central (aOR=1.43, 95% CI: 1.05-1.94) regions had higher odds of concordant prescriptions. Medications used primarily for chronic conditions had markedly higher odds of concordant prescriptions than those for acute conditions (aOR=10.94, 95% CI: 6.47-18.50). Compared with family practice, prescriptions linked to other provider types had lower odds of concordant prescriptions (aOR=0.57, 95% CI: 0.41-0.79); no significant difference was observed between family practice and obstetrics, internal medicine, or emergency medicine. Prescriptions at outpatient locations had higher odds of concordant prescriptions than those at office or clinic visits (aOR=2.83, 95% CI: 1.37-5.85). Each additional day of supply was associated with 4% lower odds of concordant prescriptions (aOR=0.96, 95% CI: 0.96-0.96) (Table 4).

**Table 4.** Multivariable regression of characteristics associated with concordant prescriptions.

| Variable | OR [95% CI] | aOR [95% CI]* | P (adj) |
| --- | --- | --- | --- |
| Age <35 | 1.25 [0.96-1.62] | 1.34 [1.02-1.75] | 0.0360 |
| <14 days past delivery | 0.40 [0.29-0.55] | 0.53 [0.38-0.73] | 0.0001 |
| Region |  |  | 0.0145 |
| South | 1 (referent) | 1 (referent) |  |
| West | 1.36 [1.02-1.80] | 1.50 [1.12-2.02] |  |
| North Central | 1.23 [0.91-1.66] | 1.43 [1.05-1.94] |  |
| Northeast | 0.98 [0.72-1.32] | 1.07 [0.78-1.46] |  |
| Unavailable | 0.39 [0.12-1.23] | 0.45 [0.13-1.57] |  |
| Maintenance |  |  | <0.0001 |
| Acute | 1 (referent) | 1 (referent) |  |
| Chronic conditions | 1.55 [1.00-2.42] | 10.94 [6.47-18.50] |  |
| Chronic and acute | 0.38 [0.30-0.48] | 1.17 [0.86-1.58] |  |
| Provider |  |  | 0.0090 |
| Family practice | 1 (referent) | 1 (referent) |  |
| OBGyn/perinatal | 0.86 [0.62-1.20] | 0.80 [0.57-1.13] |  |
| Internal medicine (NEC) | 0.66 [0.40-1.08] | 0.71 [0.43-1.17] |  |
| Emergency medicine | 1.70 [0.77-3.75] | 0.76 [0.32-1.80] |  |
| Other | 0.66 [0.48-0.91] | 0.57 [0.41-0.79] |  |
| Place of service |  |  | 0.0119 |
| Office/clinic | 1 (referent) | 1 (referent) |  |
| Outpatient | 3.70 [1.83-7.47] | 2.83 [1.37-5.85] |  |
| Other | 2.10 [1.15-3.83] | 1.53 [0.79-2.98] |  |
| Day supply (per day) | 0.97 [0.97-0.98] | 0.96 [0.96-0.96] | <0.0001 |

\*Adjustment was made for all other variables presented in the table.

## 4. Discussion

Among 96,484 new prescriptions included in the concordance analysis, more than 99% were classified as concordant while fewer than 1% were classified as discordant with published lactation compatibility classifications. Discordant prescriptions were filled later postpartum, had longer day supplies, and were more common among medications used primarily for chronic conditions and prescriptions associated with “other” provider types. Several prescription and prescriber characteristics were independently associated with concordance, including postpartum timing, chronic-condition classification, provider type, and day supply. The rarity of divergence is the primary finding, but it should not be interpreted as a direct measure of prescribing quality. It is also partly bounded by the classification itself: only 108 monographs in the 2012 database carry a category indicating potential toxicity to the breastfed infant (L4 or L5), so the proportion of prescriptions that could be discordant is small.

Drug utilization research bridges pharmacoepidemiology and health services research, and focuses on the role of medications in providing optimal care while avoiding the consequences of inappropriate drug use^13^. Within it, prescribing patterns are described through the characteristics of the prescription: the drug, the prescriber, and the patient^14^. Prescribing quality is conventionally measured through indicators that carry widespread consensus. Because there is little evidence, and therefore limited consensus, in lactational pharmacovigilance, such measures are more appropriately defined as performance indicators than quality indicators, “statistical devices for monitoring performance without any necessary inference about quality”^15^. Performance indicators permit comparison between units and are hypothesis-generating.

Comparison with other studies using these categories requires care, because results depend heavily on where the risk boundary is drawn. In a Brazilian birth cohort, 79.6% of breastfeeding women used a medication carrying some risk under Hale′s categories, but it classified L2 and L3 with L4 and L5^3^. On the boundary used here, 16.1% of women with a classifiable medication used at least one L4 or L5 medication, still higher than the 0.8% of prescriptions that were discordant in this study. Three differences likely contribute. Lutz et al. counted women rather than prescriptions, and a woman taking several medications has more opportunities to receive a discordant one. Formularies also differ: metamizole, classified L4 and widely used in Brazil, is not marketed in the United States. And that cohort captured self-medication, which claims cannot.

Concordance was lower among prescribers who see lactating patients less often. Within the lactation windows, obstetric, family practice, internal medicine, and emergency medicine prescribers wrote most of the prescriptions. Compared with family practice, concordance did not differ for obstetric, internal medicine, or emergency medicine prescriptions, but prescriptions from all other provider types had 43% lower odds of concordance. These prescribers may be less familiar with identifying concordant medications, or may treat conditions for which fewer concordant options exist.

These provider associations may also reflect differences in case mix and medication options across specialties, and the present data cannot determine whether provider familiarity, specialty-specific medication choices, or clinical necessity explain it. The pattern in Table 2 suggests how the categories are used in practice: divergence occurred almost entirely among medications for which some lactation data existed. Where data existed, prescribers appear to have been willing to use a higher-category medication, presumably because the evidence allowed a judgment about whether the risk applied to the patient; where no data existed, they did not. That is consistent with the categories serving as one input to clinical judgment rather than as a rule. Prescriptions for medications used primarily for chronic conditions had substantially higher odds of concordance than prescriptions for medications used for acute conditions. This association was considerably stronger after adjustment than in the unadjusted model and should therefore be interpreted cautiously, particularly because the analysis was based on the smaller complete-case sample. Day supply showed a more consistent association with concordance: each additional day of medication supplied was associated with a 4% decrease in the adjusted odds of concordant prescriptions. These findings identify characteristics associated with prescribing concordance but do not establish that chronic or longer-duration therapy is inherently more or less appropriate during lactation.

The rarity of divergent prescribing has an implication beyond reassurance: selection of an L4 or L5 medication appears unlikely to explain most medication-related breastfeeding discontinuation. In a recent systematic review, 110 of 112 documented cases of medication-related breastfeeding discontinuation involved medications classified as compatible, probably compatible, or presumed compatible (L1-L3); however, the review rated certainty for this outcome as very low^9^. This points to factors beyond drug-category selection, including counseling and patient perceptions, without establishing causality.

Counseling is one plausible downstream target. In a prospective Greek cohort, 57 women ceased breastfeeding because of medication intake; only 10.5% had received evidence-based counseling from a physician, and 68.4% ceased as a consequence of erroneous professional advice^4^. In a Serbian study of 1,243 breastfeeding mothers, 60.0% had been prescribed a medication while breastfeeding and 52.8% reported receiving no specific advice from their doctor about taking it^16^. These describe two distinct failures, wrong advice and absent advice, neither of which is visible in prescribing data.

Patient perceptions may compound counseling effects. In a Saudi survey of 1,831 women, 27.3% considered medication use during breastfeeding harmful^17^. Specialized pharmacist cohorts also document treatment delay driven by fear or inconsistent information^18^. Because claims record dispensed fills, prescriptions declined before reaching a pharmacy are absent from these data, and they are unlikely to be absent at random.

There is evidence that targeted counseling can change management. In 34 interprofessional pharmacist consultations for breastfeeding mothers, 18 (52.9%) involved initiating necessary medication that had previously been delayed, primarily because of fear of infant harm and conflicting recommendations; breastfeeding was resumed in seven cases following risk-benefit assessment^18^. These findings come from a small, selected consultation cohort, but they illustrate a mechanism that is not visible in claims data.

Several limitations affect interpretation of the results of the current study. Prescribing performance is determined based on lactation risk categories from a single expert reference, which may limit generalizability if expert assessment differs from common practice. No consensus alternative exists, and divergence from this classification is therefore not evidence of inappropriate prescribing. The multivariable model was restricted to the 28,732 prescriptions with complete covariate data, roughly 30% of the descriptive sample, and the loss is driven by prescriber linkage. Because this missing information was likely informative, imputation of missing data was not performed, and the complete-case estimates may be biased for the same reason. Linkage succeeds when a prescription can be matched to an outpatient encounter on the same date, which is unlikely to occur at random with respect to provider type, one of the exposures examined. Given that discordance was rare overall, sparse-data bias cannot be excluded. The model focuses on prescribing performance based on concordance in the context of infant safety rather than maternal health, potentially overlooking important aspects of medication prescribing that impact the well-being of lactating mothers. The absence of a comparison group of non-lactating women means that the method cannot account for the potential withholding of necessary prescriptions due to lactation. Finally, lactation status was inferred from surrogate indicators rather than directly recorded, and that phenotype has not been validated against a chart-confirmed standard^10^. These data describe prescriptions dispensed between 2012 and 2014 and predate the ICD-10 transition; prescribing practice has changed since. Findings are generalizable only to commercially insured individuals.

Prescribing to lactating women in a national commercially insured population rarely diverged from the 2012 Hale compatibility categories. Prescription and prescriber characteristics were associated with concordance, but associations in the full descriptive sample and the complete-case model did not always agree, particularly for postpartum timing and chronic-condition classification. Longer day supply and provider type remained informative in the adjusted model. The wide range of prescribers involved supports the conclusion that lactation pharmacology education should extend beyond obstetrics. Because concordance captures only one dimension of medication decision-making, future work should evaluate counseling, maternal treatment adequacy, and breastfeeding outcomes alongside drug selection.

## Acknowledgment

**Acknowledgments**

The authors thank Joseph Bocchino, EdD, and Anthony Scialli, MD, for their guidance on the doctoral work from which this analysis derives.

## AI statement

The authors acknowledge the use of Claude (Anthropic) to assist with manuscript preparation. The authors reviewed and verified all AI-assisted output, including numerical values and references, and take full responsibility for the content of this article.

## Author contributions

CRediT: Conceptualization: KK; Data curation: KK; Formal Analysis: KK, DA; Funding acquisition; Methodology: KK, DA; Supervision: DA; Visualization: KK; Writing – original draft: KK; Writing – review & editing: KK, DA

## Statements and Declarations

### Ethical Considerations

Deidentified patient-level data. The Office of Human Research at The George Washington University determined that this study did not constitute human subjects research and did not require Institutional Review Board review. The analysis used fully deidentified data from the Merative MarketScan Commercial Claims and Encounters Database.

### Consent to Participate

Not applicable Consent for Publication. Not applicable

### Declaration of Conflicting Interest

K.K. is Executive Director of the InfantRisk Center, Texas Tech University Health Sciences Center, and is a current co-author of Hale’s Medications and Mothers’ Milk, the reference from which the lactation risk categories used as the outcome variable in this analysis are drawn. The 2012 and 2014 editions applied here, selected to correspond with the study period, predate that authorship. K.K. had no role in assigning the lactation risk categories used in this analysis. D.A. declares no competing interests.

### Prior publication

This work derives from the corresponding author′s doctoral dissertation, The George Washington University, 2024. A companion analysis of the same cohort has been submitted to this journal.

## Funding Statement

### Funding

This research received no specific grant from any funding agency in the public, commercial, or not-for-profit sectors. Data were obtained through the Merative MarketScan Dissertation Support Program.

## Data Statement

### Data license and ownership (REQUIRED VERBATIM — do not edit)

Certain data used in this study were supplied by International Business Machines Corporation as part of one or more Merative MarketScan Research Databases. Any analysis, interpretation, or conclusion based on these data is solely that of the authors and not International Business Machines Corporation.

### Data availability

The MarketScan data underlying this analysis are licensed from Merative and cannot be redistributed by the authors. The lactation phenotype and complete code lists are published with the companion analysis. No additional data were generated for this study.

## Notes

### Author Declarations

The Office of Human Research of The George Washington University determined that this study did not constitute human subjects research and did not require Institutional Review Board review, as the analysis used fully deidentified data from the Merative MarketScan Commercial Claims and Encounters Database.

